# Sport Specialization, Injury and Illness: A Prospective Study of Elite Female Adolescent Soccer Players

**DOI:** 10.1101/2024.09.23.24314149

**Authors:** Andrew Watson, Kristin Haraldsdottir

## Abstract

**Context:** Although considerable cross-sectional evidence exists regarding the association between sport specialization and injury, sport-specific, prospective research is lacking. Similarly, sport specialization is associated with impairments in sleep and subjective well-being in youth athletes, but it is unknown whether this increases the risk of illness.

**Objective:** To determine the relationship between sport specialization status and in-season injury and illness risk in elite female adolescent soccer players.

**Design:** Prospective Cohort Study

**Setting:** Pre-season evaluation of maximal aerobic capacity (VO_2max_) and sport specialization, and in-season self-reporting of daily training load (session-rating of perceived exertion), injury and illness resulting in lost sport participation time.

**Participants:** 80 female youth soccer players (ages 13-18) from a local youth soccer organization.

**Main outcome measures:** Sport specialization status, injury and illness incidence during two 4-month competitive soccer seasons. Athletes were considered specialized if they participated in soccer exclusively versus multiple sports during the year. Mixed effects logistic regression models were used to individual injury and illness (yes/no) during the study period, using sport specialization, age, and training load as fixed effects and individual athlete as a random effect.

**Results:** Specialized (n=46) athletes did not differ from non-specialized (n=34) athletes with respect to age, preseason physical activity, VO_2max_ or in-season training load (all p>0.05). No difference was seen in the proportion of individuals from each group that reported an in-season injury (specialized = 19% v 17%, p=0.83) or illness (40% v 38%, p=0.82). After adjusting for age and training load, the individual injuries (OR= 0.86 [0.26, 2.8], p=0.81) and illnesses were not significantly predicted by specialization (OR= 1.06 [0.45, 2.5], p=0.89).

**Conclusions:** After adjusting for age and training load, sport specialization status was not associated with in-season injury or illness risk in elite female soccer players.

**Key points:**

- Prior research regarding the associations between sport specialization and health outcomes has been primarily cross-sectional and failed to account for the confounding role of training load.
- Among elite adolescent female soccer players with similar pre-season physical activity levels in-season training loads, sport specialization was not associated with injury or illness incidence across two competitive seasons.

---

The association between sport specialization and overuse injury has been widely reported among youth athletes. The majority of this research has been cross-sectional, however, and typically failed to account for the potentially confounding role of training load. Specialized athletes may be subjected to higher TL than non-specialized athletes, and increases in TL have been associated with an increased risk of injury.^1^ In a prospective study among high school athletes from multiple sports, after controlling for in-season competition volume, McGuine et al. found that high school athletes reporting moderate and high levels of sport specialization had an increased risk of in-season lower extremity injury compared to athletes with low levels of specialization. Notably, this was entirely due to the association of specialization with chronic lower extremity injuries, as no association was identified with acute lower extremity injuries.

Although it has been suggested that the relationship between specialization and injury is likely sport-specific, little prospective research has attempted to evaluate these relationships. In a single prospective study of youth basketball players, Post et al. found no statistically significant association between sport specialization and injury incidence in a prospective study of youth basketball athletes. Notably, Post et al. had previously identified a significant association between specialization and injury in a cross-sectional study of youth basketball athletes, suggesting that relationships from retrospective, cross-sectional research may not translate into sport-specific prospective injury risk.^2^ It remains unclear whether the associations between specialization and injury risk differ across sports that may have different levels of specialization, different physical demands, and different types of injuries.

Similarly, several guidelines and policy statements have suggested that athletes who choose to specialize in a single sport at an early age may be at risk of increased stress, overtraining, and burnout.^3–6^ A prior prospective study found that specialized female youth athletes demonstrated greater impairments in fatigue, mood, stress and sleep quality during the season compared to their non-specialized counterparts, even after adjusting for training load (TL).^7^ In adult athletes, higher volumes of exercise have been shown to be associated with an increased susceptibility to viral infections,^8,9^ and increased weekly TL is associated with an increased illness risk.^9,10^ A study of 15-18 year old male soccer players found that higher training duration was predictive of in-season illness,^11^ and we have previously found that higher weekly and monthly TL was predictive of in-season illness risk among adolescent female soccer players.^12^ Poor sleep and increased psychosocial stress have been suggested as risk factors for illness in athletes,^11,13,14^ but it is unknown whether youth athletes who specialize in one sport are at an increased risk of in-season illness compared to youth athletes who participate in other sports during the off-season.

In 2019, the American Medical Society for Sports Medicine hosted a research summit on youth early sports specialization, identifying a need for prospective, sport-specific research to evaluate the association between sport specialization and outcomes that controls for the confounding effects of training load. Unfortunately, since that time, well-controlled, sport-specific, prospective research in this area has been minimal. Despite the popularity of youth soccer throughout the world, however, we are not aware of research since that time which has prospectively evaluated the relationship between sport specialization, injury and illness in youth soccer athletes. In a single cross-sectional study of male youth soccer athletes, specialization was not found to be associated with an increased risk of injury,^15^ but no prior research has been conducted in female youth soccer athletes. Therefore, the purpose of this study was to determine the relationship between sport specialization and in-season injury and illness among female youth soccer players, while adjusting for the important influence of TL.

## METHODS

### Study Design

All procedures performed in this study were approved by the Institutional Review Board of the University of Wisconsin-Madison. Female athletes ages 13-18 were recruited through participation in a local youth soccer organization. Interested participants were asked to provide information regarding medications or significant cardiac or pulmonary disease that could impact cardiovascular function or performance (stimulants, congenital cardiac disease, uncontrolled hypertension, uncontrolled asthma, e.g.). Written, informed assent was provided by minor participants, and written, informed consent was provided by adult participants and parents of minor participants.

Prior to the start of two consecutive, 13-week fall club soccer seasons, participants completed a questionnaire regarding demographic information, sport participation history, and hours of weekly moderate-vigorous physical activity during the prior 4 weeks. As all of the athletes identified soccer as their primary sport and participated in soccer exclusively during the fall club season and for more than 8 months of the year, players were considered specialized if they participated in soccer exclusively throughout the year and had previously quit another sport to focus on soccer, and non-specialized if they also participated in other sports during the remainder of the year outside of the soccer season. This is similar to the categorization reported in a previously published study of specialization, well-being and sleep.^16^

Following completion of the questionnaire, participants were familiarized with the laboratory testing equipment. Height was measured with a stadiometer to the nearest 0.25 cm and body mass with a calibrated balance beam scale to the nearest 0.1 kg. Each participant then performed a progressive, maximal exercise test on an electronically braked cycle ergometer (Racermate, Seattle, WA). Subjects were familiarized with the exercise testing equipment, after which they were asked to pedal in an upright position at a cadence of 70 rpm with an initial load of 85W and incremental loads of 35W applied at 3-min intervals to the point of exhaustion, defined as the point when the pedal cadence can no longer be maintained despite strong verbal encouragement. During the test, total ventilation, oxygen consumption (VO_2_), and carbon dioxide (VCO_2_) were measured, and respiratory exchange ratio (RER; VCO_2_/VO_2_) was calculated, in a continuous, breath-by-breath manner by a metabolic cart (Cosmed, Chicago, IL). To account for variations in breath-by-breath measurement, these values were expressed as a 30-second rolling average. VO_2max_ was defined as the highest value obtained during the test, and recorded along with time to exhaustion (T_max_). VO_2max_ was expressed relative to body mass (ml/kg/min). A test was considered maximal if the athlete satisfied at least 2 of the following 3 objective criteria: (1) maximal heart rate >90% of the predicted maximal heart rate; (2) RER >1.1; (3) a plateau in oxygen consumption, defined as a change of <2 ml/kg/min in O_2_ consumption over the last 60 seconds of the test. All of the fitness testing was conducted by a primary care sports medicine physician and an exercise physiologist with several years of prior testing experience.

Immediately following all physical activity, participants provided the duration (minutes) and perceived intensity (1 to 10) of the session using an online software program (fitfor90.com), which were multiplied to yield a unitless session-rating of perceived exertion (sRPE) value as a measure of internal TL.^10,17^ Throughout each season, participants were asked to report any injuries that resulted in restriction from participation in soccer events, including the date of onset, injury location, injury type, injury mechanism (acute, chronic) and diagnosis. Similarly, athletes were asked to report any illnesses that resulted in restriction from participation, including the date of onset, predominant symptoms, and diagnosis, using the same online software program. Follow-up interviews were conducted by the study team in any instances in which the details of the injury or illness were unclear. Compliance with the completion of daily TL, injury and illness reporting was encouraged periodically throughout the study period by coaching staff. This approach has been reported previously in prior studies of the predictors of in-season injury and illness among youth soccer athletes.^18,19^

#### Statistical Analysis

Data were initially evaluated for normality using descriptive statistics and histogram analysis. Age group was determined based on birth year in accordance with the current classification by U.S. Soccer. Age, BMI, preseason physical activity, years of soccer experience, and fitness (VO_2max_, T_max_) were compared between specialization groups with Wilcoxon Rank Sum Tests and across age groups (U14, U15, U16, U17, U18) using Kruskal-Wallis tests with post-hoc pairwise Wilcoxon Rank Sum Tests adjusted for multiple comparisons. The proportion of individuals who suffered an in-season injury or illness were compared across age groups (U14, U15, U16, U17, U18) using a chi-square test. Separate mixed effects logistic regression models were used to predict 1) in-season injury (yes, no) and 2) in-season illness (yes, no) for each individual using sport specialization, age, and TL as fixed effects. Individual was included as a random effect since some participants participated in multiple seasons. Finally, separate survival analyses were conducted to compare 1) time to injury and 2) time to illness between the specialized and non-specialized athletes, with comparisons using the Likelihood Ratio Test from similarly adjusted Cox proportional hazards models. In the comparisons between specialization groups and in the mixed effects model, age at the start of the season was included as a continuous variable, rather than age group. The participants in this study represent a convenience sample of athletes from a single soccer club who were interested in participation during two consecutive years and does not represent a sample size based on an *a priori* power analysis. Significance level was determined *a priori* at the 0.05 level and all tests were 2-tailed. All statistical analyses were performed in R.^20^

## RESULTS

Eighty female youth soccer athletes (15.2 ± 1.5 years) participated in the study, with 46 specialized and 34 non-specialized athletes. 56 athletes (37 specialized) participated in the first year, while 52 (31 specialized) participated in the second year, 28 of whom participated in both years (22 specialized), resulting in 108 person-years over the two seasons. There were no differences between groups with respect to age, years of experience, pre-season physical activity, or fitness level (Table 1). The proportion of athletes categorized as specialized differed significantly across age groups, with older age groups demonstrating higher rates of specialization (Figure 1). Total in-season TL during the two seasons did not differ between specialized and non-specialized athletes (417 ± 101 v 393 ± 100 arbitrary units, p=0.31).

**Figure 1.**
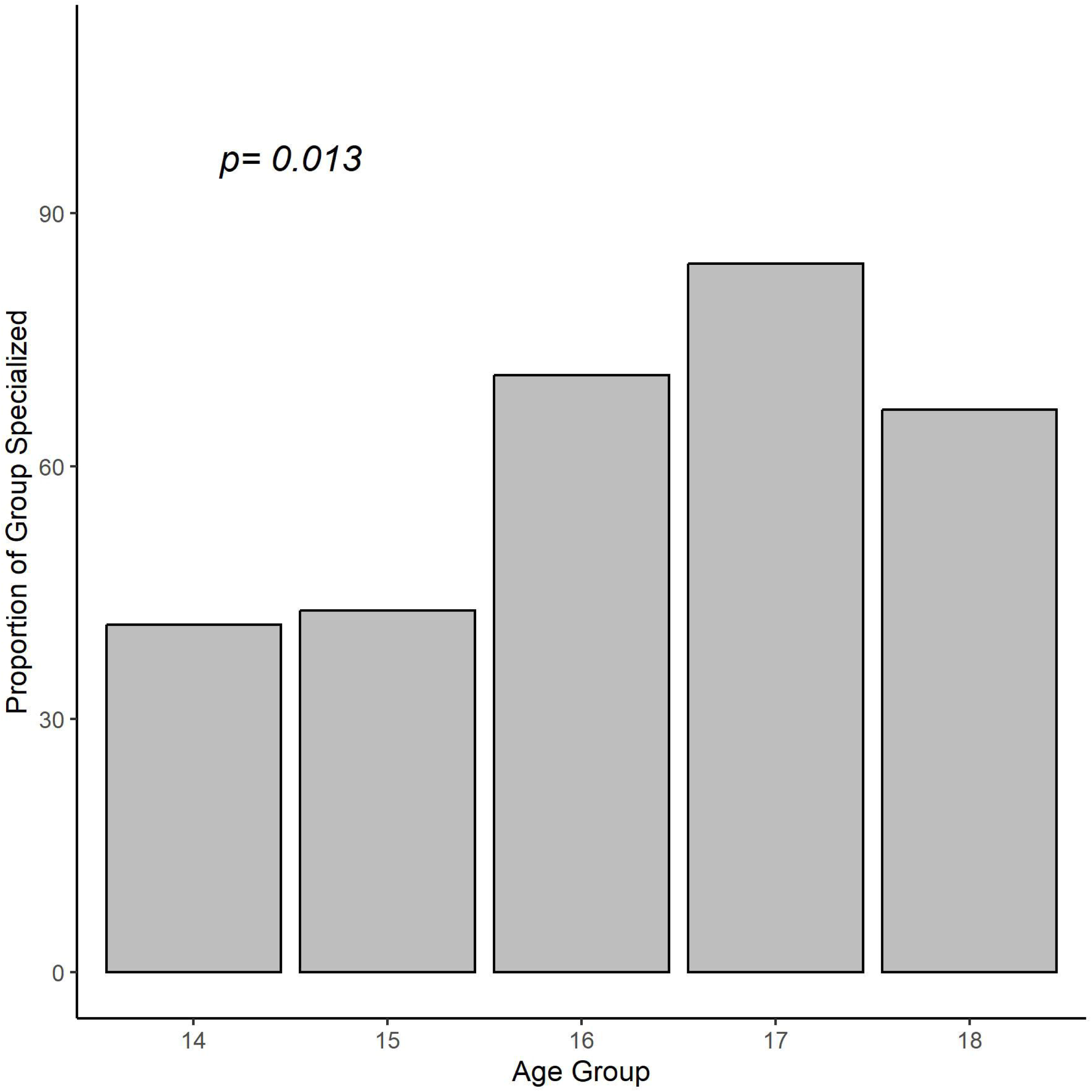
The proportion of adolescent female soccer players in each age group who were classified as specialized.

**Table 1.** Comparison of pre-season age, soccer experience, physical activity, fitness and prior injury between specialized and non-specialized female soccer athletes.

| Variable | Specialized (n=46) | Not specialized (n=34) | *p* |
| --- | --- | --- | --- |
| Age (years) | 15.21 ± 1.5 | 14.7 ± 1.7 | 0.28 |
| Soccer experience (years) | 9.4 ± 2.0 | 9.4 ± 3.4 | 0.74 |
| Pre-season PA (hours/week) | 3.3 ± 1.3 | 3.7 ± 1.2 | 0.37 |
| VO<sub>2max</sub> (ml/kg/min) | 47.2 ± 6.5 | 46.5 ± 6.1 | 0.56 |
| Time to exhaustion (min) | 14.3 ± 2.2 | 14.8 ± 2.9 | 0.48 |

Data shown as mean ± SD. BMI = body mass index; PA = hours of vigorous physical activity;
VO<sub>2max</sub> = maximal aerobic capacity.

Twenty-eight time-loss injuries were reported, of which 3 (11%) were chronic and knee, ankle and upper leg were the most common injury locations (Table 2). The proportion of athletes that suffered an in-season injury during the study period did not differ between specialized and non-specialized groups (19% v 17%, p=0.83). In the mixed effects logistic regression model, in-season injury was not independently predicted by specialization (odds ratio (OR) = 0.86 [95% CI = 0.26 – 2.8], p=0.81), training load (OR = 0.61 [0.30 – 1.23], p=0.18), or age (OR = 1.1 [0.71–1.7], p=0.68). In the survival analysis, no significant differences were identified between specialized and non-specialized athletes with respect to time to injury (p=0.16; Figure 2).

**Figure 2.**
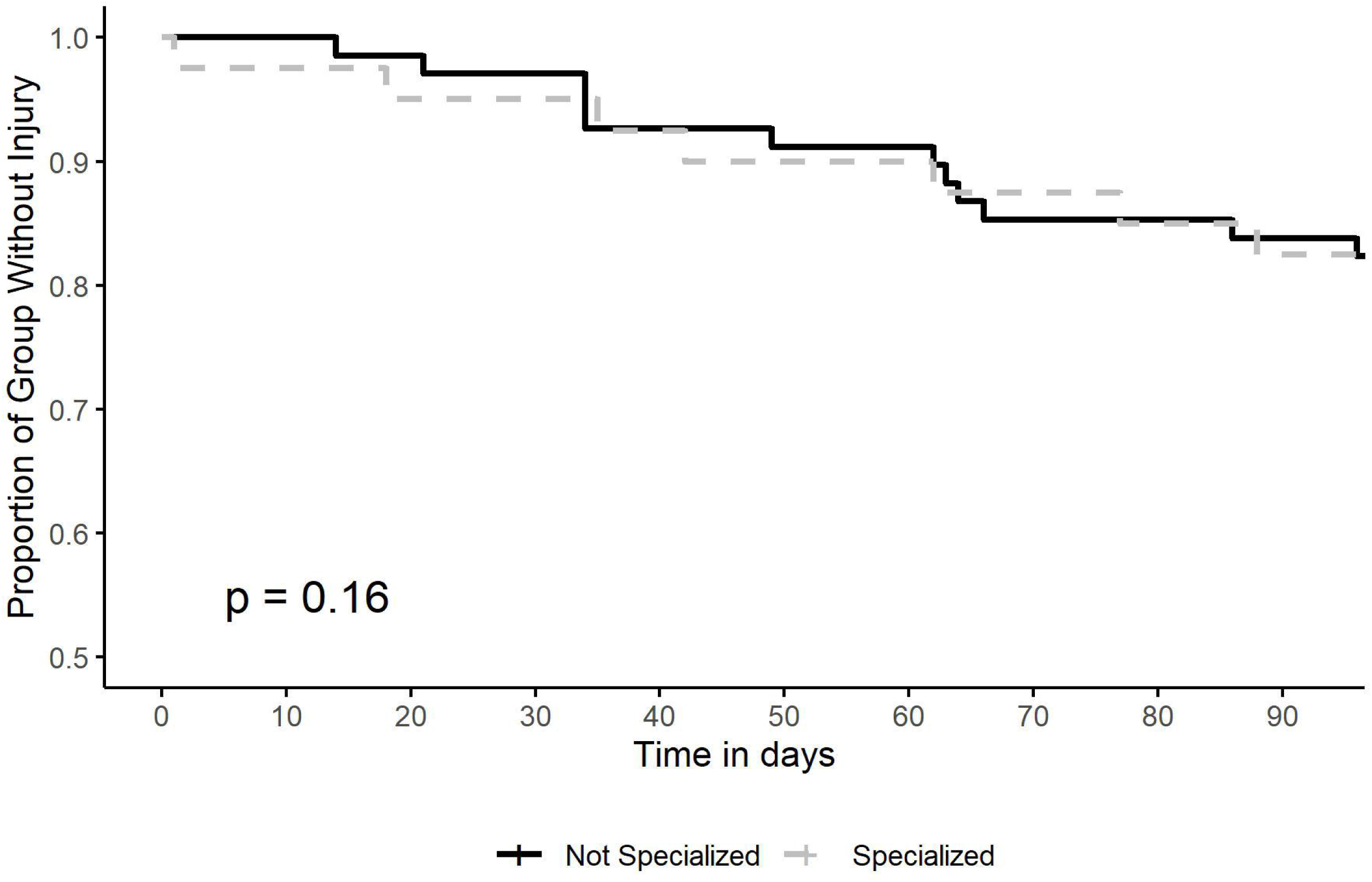
Kaplan-Meier curve showing the proportion of specialized and non-specialized female adolescent soccer athletes that remained free of injury over the course of two 4-month seasons.

**Table 2.**
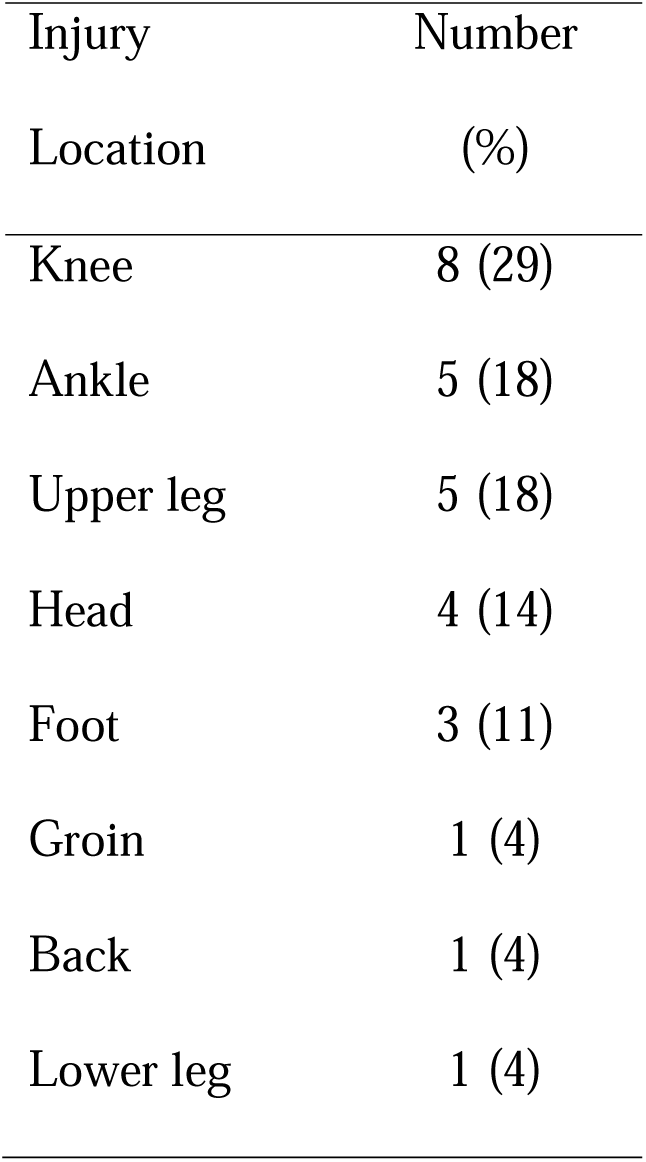
Injury locations reported over two consecutive seasons among elite female adolescent soccer players.

Fifty-nine illnesses were reported, with the most common being upper respiratory illnesses, followed by gastrointestinal illnesses (Table 3). The proportion of athletes that suffered an in-season illness during the study period did not differ between specialized and non-specialized groups (40% v 38%, p=0.82). In the mixed effects logistic regression model, in-season illness was not independently predicted by specialization (OR= 1.06 [0.45 – 2.5], p=0.89), training load (OR= 1.4 [0.92 – 2.1], p=0.12), or age (OR= 1.13 [0.83 – 1.5], p=0.44). Similarly, no significant difference was found in the survival analysis between specialized and non-specialized athletes with respect to time to illness (p= 0.81; Figure 3).

**Figure 3.**
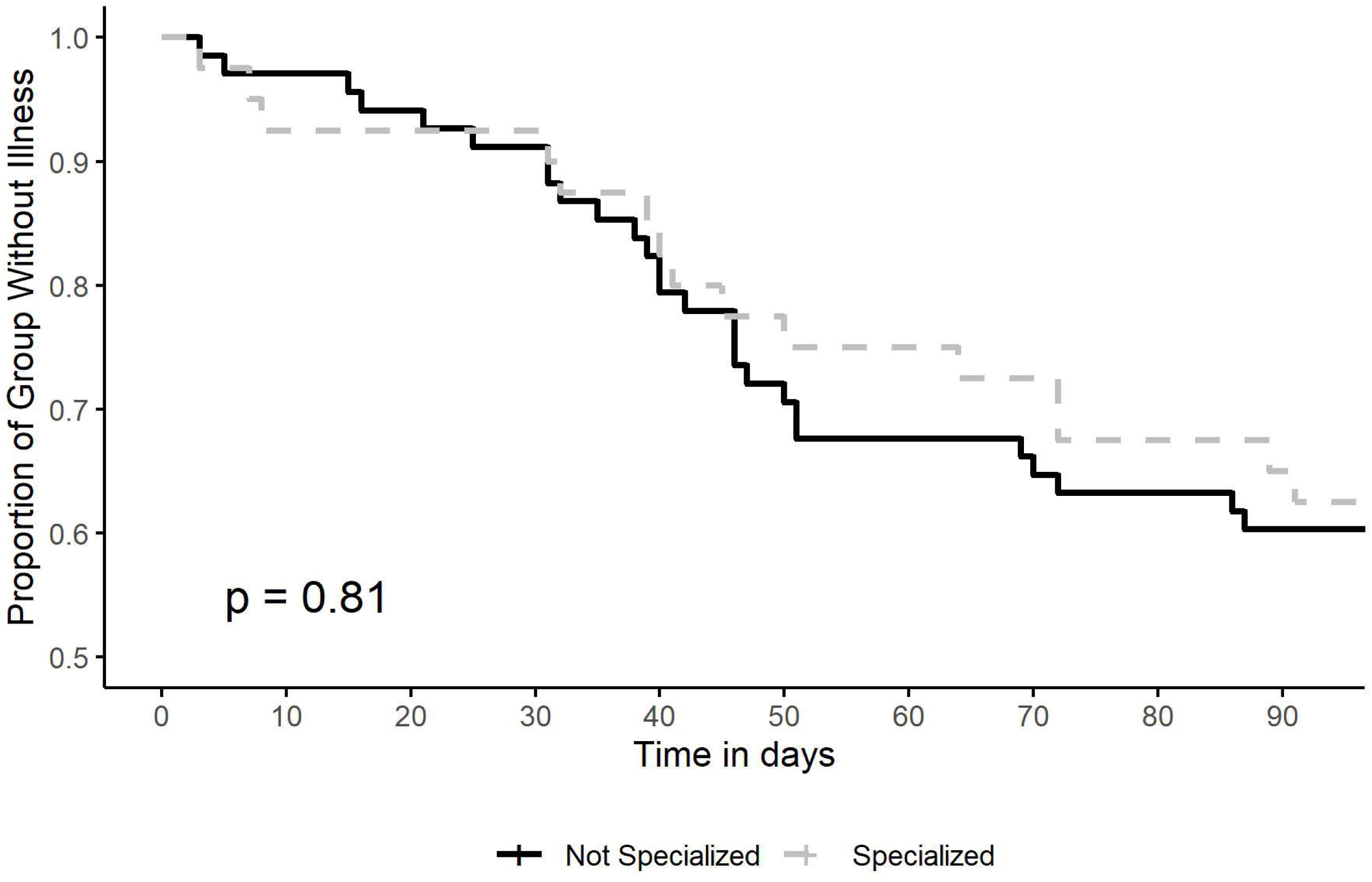
Kaplan-Meier curve showing the proportion of specialized and non-specialized female adolescent soccer athletes that remained free of illness over the course of two 4-month seasons.

**Table 3.** Types of illnesses reported over two consecutive seasons among elite female adolescent soccer players.

| Illness Type | Number (%) |
| --- | --- |
| Upper respiratory illness | 44 (73) |
| Gastrointestinal illness | 8 (13) |
| Lower respiratory illness | 4 (7) |
| Fever | 3 (5) |

## DISCUSSION

The primary finding of this study was that we found no evidence of an increased risk of in-season injury among specialized youth soccer athletes over the course of two consecutive seasons. Similarly, we found no difference in the time to injury between athletes who exclusively participate in soccer and those who participate in multiple sports. This is contrast to the cross-sectional evidence regarding the relationships between specialization and injury, as well as a large, prospective study of specialization and injury among high school athletes from various sports.^21–23^ It is, however, similar to the findings of a prior cross-sectional study of elite youth male soccer athletes, as well as a recent, prospective study of youth basketball athletes, both of which failed to identify an association between specialization and injury.^2,15^ This may suggest that the relationships between injury risk and sport specialization differ between sports, and reinforces the need for sport-specific evidence to help guide individualized decision-making for children and families regarding sport participation.

In addition, we found no increased incidence of illness among specialized athletes compared to their non-specialized teammates. Interestingly, sport specialization among female adolescent athletes has been found to be associated with impairments in sleep quality, mood, and fatigue,^7^ all of which can lead to increased illness risk.^24–26^ Given that higher incidence of illness among adolescent male soccer players has been found to be associated with increases in psychosocial stress,^11^ we sought to determine whether participating in other sports outside of the main competitive soccer season could have an influence on illness risk in elite female adolescent soccer players. Our results are similar to those recently reported in younger athletes, ages 10-14, that similarly found that sport specialization was not associated with increased illness risk.^27^ In fact, that study found that decreased sleep and lower physical activity were the primary predictors of in-season illness. Given the rich data in support of the positive long-term benefits of physical activity among youth and adolescents,^28^ our findings suggest that sport participation does not increase risk of illness in adolescent elite female soccer players, regardless of sport specialization status.

There are several variables which can potentially influence the relationships between sport specialization and health outcomes of athletes. TL is associated with injury risk, and it has been suggested that increased TL may be a predictor of in-season illness in athletes.^12,29^. Higher TL is often considered a potentially important confounder in comparisons of specialized and non-specialized athletes, as it is assumed that specialized athletes may have a higher TL which may be the true driver of adverse outcomes such as injury or illness. In fact, a recent review found that of the five included studies that attempted to evaluate the association between specialization and injury independent of training load, two studies found that the association was no longer significant, while three found that the relationship persisted. In order to account for this, we specifically recruited a group of athletes from within a single organization that would have comparable in-season TL, identified the specialized and non-specialized players, and adjusted for TL within our statistical models. As expected, we did find very similar in-season TL between the groups, but even after accounting for any differences we did not find any association between specialization and either injuries or illnesses during the season.

A rapid change in physical activity or training load may also impact risk of both injury and illness,^25^ where specialized athletes could potentially be at greater risk if their pre-season physical activity levels were significantly lower than the athletes participating in other sports. Similarly, we have previously found that lower levels of preseason fitness are associated with an increased risk of subsequent in-season injury and illness in youth soccer athletes.^19^ In the present study, however, we found no significant difference in VO_2max_ or pre-season vigorous physical activity between the specialized and non-specialized groups, suggesting that specialization status did not influence aerobic fitness or activity level in the soccer off-season.

This study has several limitations. Without randomization of individuals to specialization groups, it is not possible to determine whether between-group differences can be attributed to specialization itself, or some other coincident attributes of the groups that could be associated with specialization. Compliance with data collection of TL was ∼80% and as a result there was data missing. We did not identify any obvious relationship between compliance and date, specific individuals, or specialization group, and consequently chose to analyze the raw data without imputation, similar to prior work using a similar data collection mechanism.^7,16,18,19^ This study also used a commercially available software program rather than a previously utilized questionnaire regarding injury and illness reporting. Nonetheless, this approach has been used previously to identify relationships between TL, subjective well-being, injury and illness,^12^ and was felt to represent a more cost-effective and feasible method to collect large amounts of daily, individual data over a significant period of time than traditional questionnaires. A proportion of the participants participated in both years of the study, of whom a greater number were from the specialized group. While it is possible that this could skew the results, we have attempted to account for this by utilizing a mixed effects regression model to evaluate the relationships between specialization, injury and illness and included each individual as a random effect. As above, the sample size in the study was based on a convenience sample rather than an *a priori* power analysis, which could result in a failure to identify significant differences in group comparisons. Lastly, this study included only adolescent female soccer players from a single club and a single sport and may not be generalizable to other populations.

In conclusion, we did not find any association between sport specialization and in-season injury or in-season illness incidence among adolescent female soccer players. Similarly, we found no difference in the time to injury or illness between specialized and non-specialized athletes. This suggests that the relationships between sport specialization and health outcomes may be sport-specific, and further prospective research is warranted to determine whether this relationship exists in other populations of youth athletes.

## Data Availability

Data are available upon reasonable request to the authors

